# Convalescent plasma as potential therapy for severe COVID-19 pneumonia

**DOI:** 10.1101/2020.09.01.20184390

**Authors:** Ricardo Valentini, José Fernández, Dardo Riveros, Fernando Pálizas, Jorge Solimano, Pablo Saúl, Juan Medina, Viviana Falasco, María Laura Dupont, Julia Laviano, Florencia Fornillo, Daniela Maymó, Daniel Gotta, Alfredo Martinez, Pablo Bonvehí, Juan Dupont

**Affiliations:** Departamento de Medicina, CEMIC; Medicina Transfusional, CEMIC; Sección Hematología, CEMIC; Terapia Intensiva, Sanatorio Güemes; Servicio de Clínica Médica, Hospital Pedro Fiorito; Servicio de Infectología, Policlínico de Unión Obrera Metalúrgica; Terapia Intensiva, Sanatorio Itoiz; Instituto Universitario, CEMIC; Departamento de Análisis Clínicos, CEMIC; Sección Infectología, CEMIC

**Author notes:** Corresponding Author: Ricardo Valentini.

**Keywords:** COVID-19 convalescent plasma treatment, severe acute respiratory syndrome coronavirus 2, COVID-19 pandemic

## Abstract

At the beginning of the COVID-19 pandemic, there was high mortality and a lack of effective treatment for critically ill patients. Build on the experience in argentine hemorrhagic fever with convalescent plasma, we incorporated 90 patients into a multicenter study, and 87 were evaluable. We collected 397 donations from 278 convalescent donors. Patients received plasma with an IgG concentration of 0.7-0.8 (measured by Abbott chemiluminescence) for every 10 kg of body weight. Survival during the first 28 days was the primary objective. 77% were male, age 54 ± 15.6 y/o (range 27-85); body mass index 29.7 ± 4,4; hypertension 39% and diabetes 20%; 19.5% had an immunosuppression condition; 23% were healthcare workers. Plasma was administered to 55 patients (63%) on spontaneous breathing with oxygen supplementation (mainly oxygen mask with reservoir bag in 80%), and 32 patients (37%) were infused on mechanical ventilation. The 28-day survival rate was 80%, with 91% in patients infused on spontaneous breathing and 63% in those infused on mechanical ventilation (p = 0.0002). There was a significant improvement in the WHO pneumonia clinical scale at 7 and 14 days, and in PaO_2_ / FiO_2_, ferritin and LDH, in the week post-infusion. We observed an episode of circulatory volume overload and a febrile reaction, both mild. Convalescent plasma infusions are feasible, safe, and potentially effective, especially before requiring mechanical ventilation, and are an attractive clinical option for treating severe forms of COVID-19 until other effective therapies become available.

---

The outbreak of severe acute respiratory syndrome coronavirus 2 (SARS-CoV-2), which was originated in Wuhan, China, has become a major concern worldwide ^1^. Pneumonia induced by SARS-CoV-2 is the leading cause of death of severely ill patients and despite of intensive research, no proven therapy for severe respiratory disease has been yet described better than conventional support ^2^. Antibiotics, antivirals, antiparasitic and a variety of anti-inflammatory drugs and biologicals, are included in the support therapy, although with potential side effects. The identification of alternative strategies is needed in the setting of severely ill patients. The recent low dose (6mg for 10 days) dexamethasone arm of the Recovery Trial has emerged as a concrete evidence for the use of the drug in patients with severe pneumonia ^3^.

Convalescent plasm has been used to transfer passive immunity in viral diseases. A double-blind controlled study was conducted in Argentina, between 1974 and 1978, in patients with Argentine hemorrhagic fever (AHF. Immune plasma (vs. normal plasma), was infused during the first week from the onset of symptoms. From 188 subjects included in the trial, fatality rate among the cases treated with normal plasma was 16.5%, while it was 1.1% with convalescent plasma ^4^.

This experience in AHF and in other epidemics like “Spanish” flu, SARS, Ebola, measles and H1N1, leaded us and others to design trials with convalescent plasma in SARS-CoV-2 infected patients. Several small observational series of cases, published early 2020, suggested that this is a potential effective strategy for severely ill patients ^5^. Safety was addressed by an extensive experience of 20,000 convalescent plasma infusions showing less than 1% serious side effects^6^. The Expanded Access Program for COVId-19 announced that 57.630 patients received convalescent plasma with similar safety profile ^7^.

We performed the present study on severe ill COVID-19 patients to provide data on clinical characteristics and outcome after plasma therapy for COVID-19. We built a network of 25 public and private hospitals of Buenos Aires urban and suburban area to evaluate feasibility, safety and potential efficacy in severe COVID-19 patients.

## Methods

### Research design and ethics

The present study was conducted at CEMIC (Centro de Educación Médica e Investigaciones Clínicas) that is university hospital in the metropolitan area of Buenos Aires. We designed a multicenter open label trial. Twenty-five public and private hospitals initially enrolled patients in the study. Protocols of donation and infusion to patients was designed by investigators at CEMIC and both approved by the Institutional Review Board. Network institutions shared the CEMIC protocol and submitted it to their local IRB, or institutional authorities for approval. The protocol was also submitted to the National Blood Authority (Health Public Ministry) and is registered in the PRIISA.BA, a public research registry of the Government of the City of Buenos Aires. Patients of all institutions received plasma units from donators of CEMIC and was devoid of any financial charge.

### Donation

Plasma donators were obtained from community volunteers that had proven COVID-19, and they were tested negative viral RNA in nasopharyngeal swabs at the time of donation. A call center run by teaching physicians and students of the medical career at the University Institute CEMIC, screened, qualified, and then scheduled donors at the Transfusion Medicine Unit at the CEMIC Hospital. A written informed consent was obtained from each donor by a study authorized physician. Plasma donation was made by conventional whole blood donation, centrifugation and autologous red blood cell reinfusion. An average of 300 ml of plasma was obtained from each donation. A total of 397 donations of plasma units were obtained of 278 donors. There was an average of 1.4 donations per donor throughout the study. Previously pregnant women were studied for HLA antibodies. Donation program begun April 8, 2020. Data were drawn at the cut off July 27^th^, 2020.

### Antibody testing in plasma donors and selection of units to be transfused

The SARS-COV-2 IgG antibody test was performed on donor serum samples using the Architect Plus i2000sr Analyzer (Abbott, Illinois, USA) and the CMIA SARS-COV-2 IgG kit. It is a chemiluminescent microparticle immunoassay for the detection of IgG in human serum or plasma against the SARS nucleoprotein CoV-2. Index values obtained from the collected plasmas ranged between 0 and 10 (mean 5.7), those higher than 3 were arbitrarily considered useful, due to their potential neutralizing capacity on the virus ^8^. Patients received the required volume of antibody plasma to achieve a dose of 0.7-0.8 / 10 kg body weight.

### Patient eligibility

Adult patients ≥18, and non-pregnant women were eligible if they had severe or critical COVID-19 disease with ≤ 10 days from the onset of symptoms or ≤ 7 days on mechanical ventilation. Severe disease was defined as one or more of the following: blood oxygen saturation ≤ 94% on supplemental oxygen by nasal cannula at least 3 L/min, non-rebreathing mask (NRO2-mask) or on noninvasive ventilation; and pulmonary infiltrates with >50% increase within 24 to 48 hours in chest-X-ray or chest CT. Life-threatening disease was defined as one or more of the following: respiratory failure on mechanical ventilation with PaO_2_ / FiO_2_ less than 300 mm Hg, septic shock, and/or multiple organ dysfunction.

### Patient enrollment

Once IRB approved the protocol physicians taking care of severely and critically ill patients in intensive care units at CEMIC and from the network, shared clinical features and images and decided enrollment. A written informed consent was obtained from the patient or a legally authorized representative. ABO and Rh typing and body weight were routinely obtained, and a suitable ABO compatible unit of plasma was delivered. The infusion rate was 150 ml/hr or at a slower rate if cardiac overload risk was suspected. Clinical data for patients were obtained from the hospital electronic medical records or a shared spreadsheet from the network institutions. First infusion was made April 18, 2020.

Data collected included: demographic characteristics, comorbidities, symptoms from baseline to plasma infusion, length of hospital stay and mechanical ventilation before plasma infusion; ventilatory parameters if they were on mechanical ventilation and serum biomarkers at the inclusion in the study and then at 3, 5 and 7 days after the infusion (C-reactive protein, D-dimer, ferritin and LDH).

The primary endpoint was survival rate at 28 days after plasma infusion. Clinical efficacy was evaluated according to the WHO scale prior to infusion, 7 and 14 days after plasma therapy. Ventilatory status was evaluated at days 1, 3, 7, and 14 and weekly up to discharge or death. The WHO clinical progression scale contains 10 variables: 0: not infected, no viral RNA detected; 1: asymptomatic, viral RNA detected; 2: symptomatic, independent of assistance; 3: symptomatic, assistance needed; 4: hospitalized, without oxygen therapy; 5: hospitalized, oxygen by mask or nasal cannula; 6: hospitalized, oxygen due to NIV or high flow; 7: intubation and mechanical ventilation, PO2 / FiO2 ≥ 150 or SaO2 / FiO2 ≥ 200; 8: mechanical ventilation, PO2 / FiO2 <150 (SaO2 / FiO2 <200) or vasopressors; 9: mechanical ventilation, PO2 / FiO2 <150 and vasopressors, dialysis or ECMO; 10: death ^9^.

To determine the potential clinical efficacy, a response to plasma infusion was considered as being alive for 28 days, with a hospitalization time ≤ 21 days or a length of stay on mechanical ventilation ≤ 14 days, compared to patients who died or, although they have survived, presented ≥ 21 days of hospitalization time, or ≥ 14 days on mechanical ventilation. Inflammatory parameters (ferritin, LDH and D-Dimer) and arterial oxygenation within the first week post-infusion were also compared between groups.

### Statistics

Descriptive variables are expressed as means ± SD, or medians and interquartile ranges (IQR) for continuous variables with normal and non-normal distribution, respectively. Paired comparisons were made using the Wilcoxon signed rank test. To compare proportions, the χ2 or Fisher’s exact test was applied, and the Friedman test was used for the paired comparison between groups with non-parametric variables and then, Bonferroni correction test was applied. A P value less than 0.05 was considered significant. The Kaplan-Meier method was used to estimate survival.

## Results

### Population

Since April 18^th^ up to July 27^th^, 90 patients were infused with plasma from COVID-19 convalescent donors. Three of them had comorbid conditions that prevented progress in therapeutic efforts immediately after the infusion and for this reason were excluded, leaving 87 patients for analysis. Among the demographic data, there was a predominance of male patients, 77% with a male / female ratio of 3.4 / 1; the median age was 54 years (± 15.6, range 27-85). The mean body mass index (BMI) was 29.7 (± 4.4, range 25-37). The most frequent comorbidities were arterial hypertension (38.7%), type II diabetes (20.5%) and morbid obesity (BMI ≥35) in 12 (13.8%) patients. 19.5% had some condition considered immunosuppression (organ transplant and autoimmune diseases). Cardiopulmonary disease was present in 19.5% (COPD, asthma, heart failure, and coronary disease), and 5 patients (5.8%) had oncological pathology. Twenty patients (23%) were healthcare workers (Table 1a).

**Table 1a.** Baseline characteristics of patients with COVID-19 pneumonia at the time of plasma treatment.

|  |  |
| --- | --- |
| **Number of Patients** | **90** |
| Non eligible | 3 |
| **Demographic Data** |  |
| Mean age, years ± DE (range) | 54.7 ± 15.6 (27-85) |
| Male sex (%) | 67 (77) |
| Mean Body mass index ± DE | 29.7 ± 4.4 |
| Health workers, n (%) | 20 (23) |
| **Comorbidities** | **n (%)** |
| Arterial Hypertension | 34 (38,6) |
| Type II Diabetes | 18 (20,5) |
| Morbid Obesity, BMI ≥35 | 12 (13,8) |
| COPD/Asthma | 9 (10,3) |
| Cardiac disease | 8 (9,1) |
| Immunosuppressed | 7 (8) |
| Solid Tumors and Hematologic malignancies | 5 (5,8) |
| Chronic renal failure | 3 (3,4) |
| Solid organ Transplant (liver, kidney) | 2 (2,2) |

Most of the patients were in the intensive care unit at the time of the plasma infusion (82%) and in 16 cases the plasma was infused in the general ward or high dependency unit. Thirty-two (37%) patients were transfused on invasive mechanical ventilation, and 55 (63%) patients received plasma while on supplemental O_2_ (by nasal cannula in 20% of subjects or by NRO-mask in 80%). In mechanically ventilated patients, the median PaO_2_ / FiO_2_ at the time of inclusion was 149 mm Hg (IQR 109-185), with 24% of patients meeting Berlin criteria for severe acute respiratory distress syndrome (ARDS); ventilation in prone position was applied in 22 of these patients (69%) and hemodialysis in 9 patients (12%).

The patients received treatments according to the therapeutic recommendations that emerged during the pandemic. Initially, patients received lopinavir-ritonavir (22 patients); 7 hydroxychloroquine with or without azithromycin and none of the last 32 included patients, received these drugs. Corticosteroids were used in 42 cases, mostly dexamethasone. Antithrombotic prophylaxis with enoxaparin was performed in accordance with institutional recommendations.

### Infusion and safety of convalescent plasma

Plasma was administered at a median of three days after hospital admission, corresponding to eight days from the onset of symptoms. In patients who received the infusion with mechanical ventilation, the time from intubation to infusion had a median of 1 day (Table 1b). All patients received 300-600 ml of plasma with CMIA IgG values between 0.7 and 0.8 per 10 kg of body weight. Median of IgG antibodies in the infusions bags was 6.5 (RIC 4-11.5). Antibodies concentration were unknown in the first five infusions. Retrospectively two had 0 and 15 index value (out of the prefixed value). In 29 patients (33.3%) the infusion of a similar dose was repeated after 48-72 h according to clinical evolution and/or the persistence on viral RNA. There were no serious adverse events (grade 3-4) attributed to plasma transfusion within 24 hours of transfusion. There was a related febrile episode and one probably related to cardiac volume overload, both mild events which did not require stop the infusion.

**Table 1b.** Characteristics of the clinical condition and concomitant supportive therapies of patients treated with plasma.

|  |  |
| --- | --- |
| **Evaluable infused patients (n)** | **87** |
| Infused with O<sub>2</sub> support n (%) | 55 (63,2) |
| Oxygen mask with reservoir bag n (%) | 44 (80) |
| Nasal cannula n (%) | 11 (20) |
| Infused under mechanical ventilation n (%) | 32 (36,8) |
| PO<sub>2</sub> / FiO<sub>2</sub> (median-IQR) | 149 (109-185) mm Hg |
| Prone position n (%) | 22 (69%) |
| hemodialysis n (%) | 9 (12%). |
| **Time to plasma infusion** | **median (IQR 20-75)** |
| From symptoms onset | 8 (5-10) |
| From Hospitalization | 3 (1-6) |
| From intubation and mechanical ventilation | 1 (0-2) |

### Clinical Outcome

Global survival at 28 days after infusion was 80%; 91% for patients who were infused with O_2_ support, and 63% for those treated with invasive mechanical ventilation (p = 0.0002) (fig. 1). The intubation rate for patients with respiratory failure and O_2_ support was 25%.

**Figure 1.**
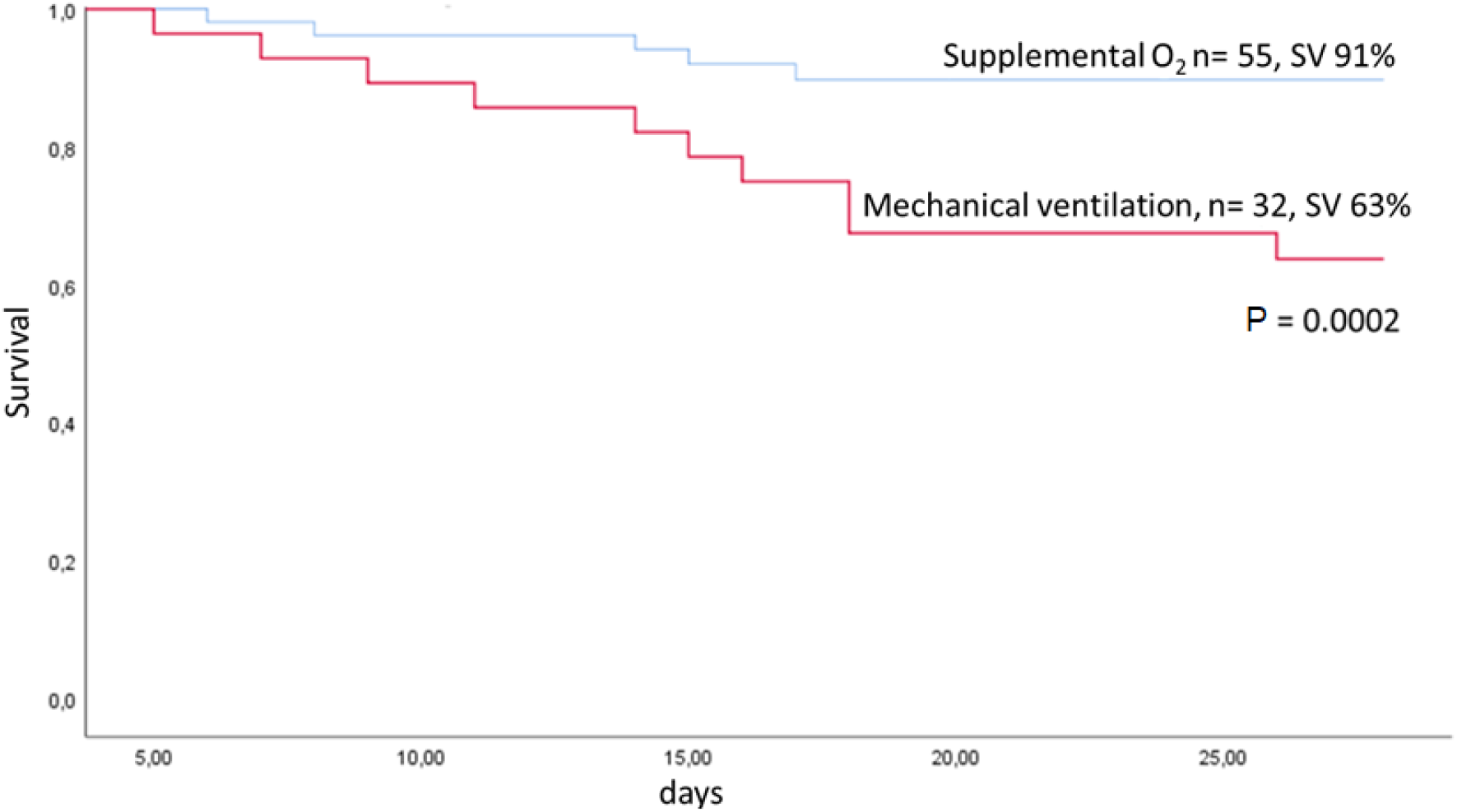
Survival of patients infused with convalescent plasma under supplemental 0_2_ compared to mechanical ventilation

The 10-point WHO ordinal clinical scale score improved significantly at 7 and 14 days after infusion by at least 1 and 2 points respectively (fig. 2). In 72% of the patients a better score was observed in the evaluation at day 7 and in 64% at day 14 (p˂ 0.001).

**Figure 2.**
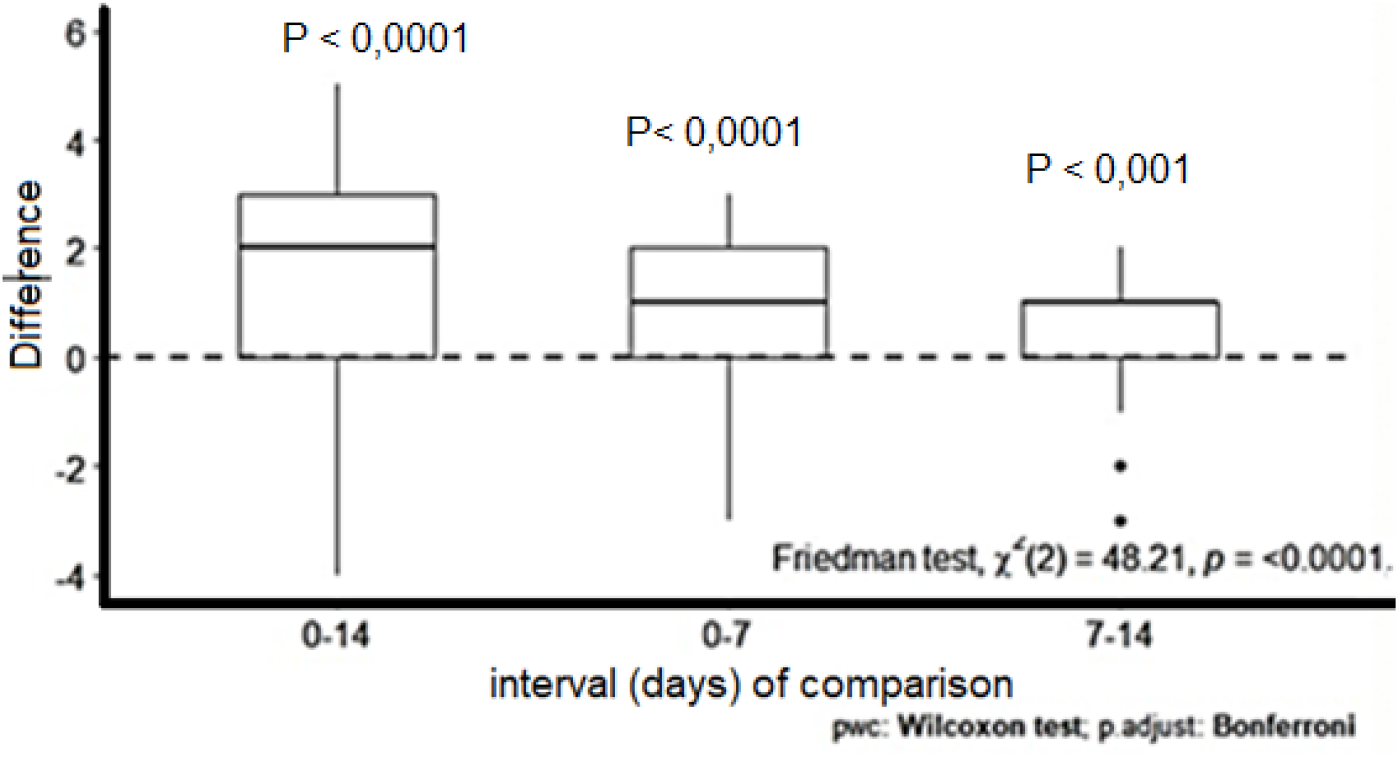
Evolution of the WHO pneumonia severity scale

Among patients that survived at day 28, 60% had a length of stay on mechanical ventilation ≤ 14 days and / or a length of hospitalization ≤ 21 days. An improvement was observed during the first week after plasma infusion in respiratory parameters evaluated by PaO_2_ / FiO_2_ and in inflammatory parameters such as LDH and ferritin levels, without significant differences when D-Dimer was considered (fig. 3)

**Figure 3.**
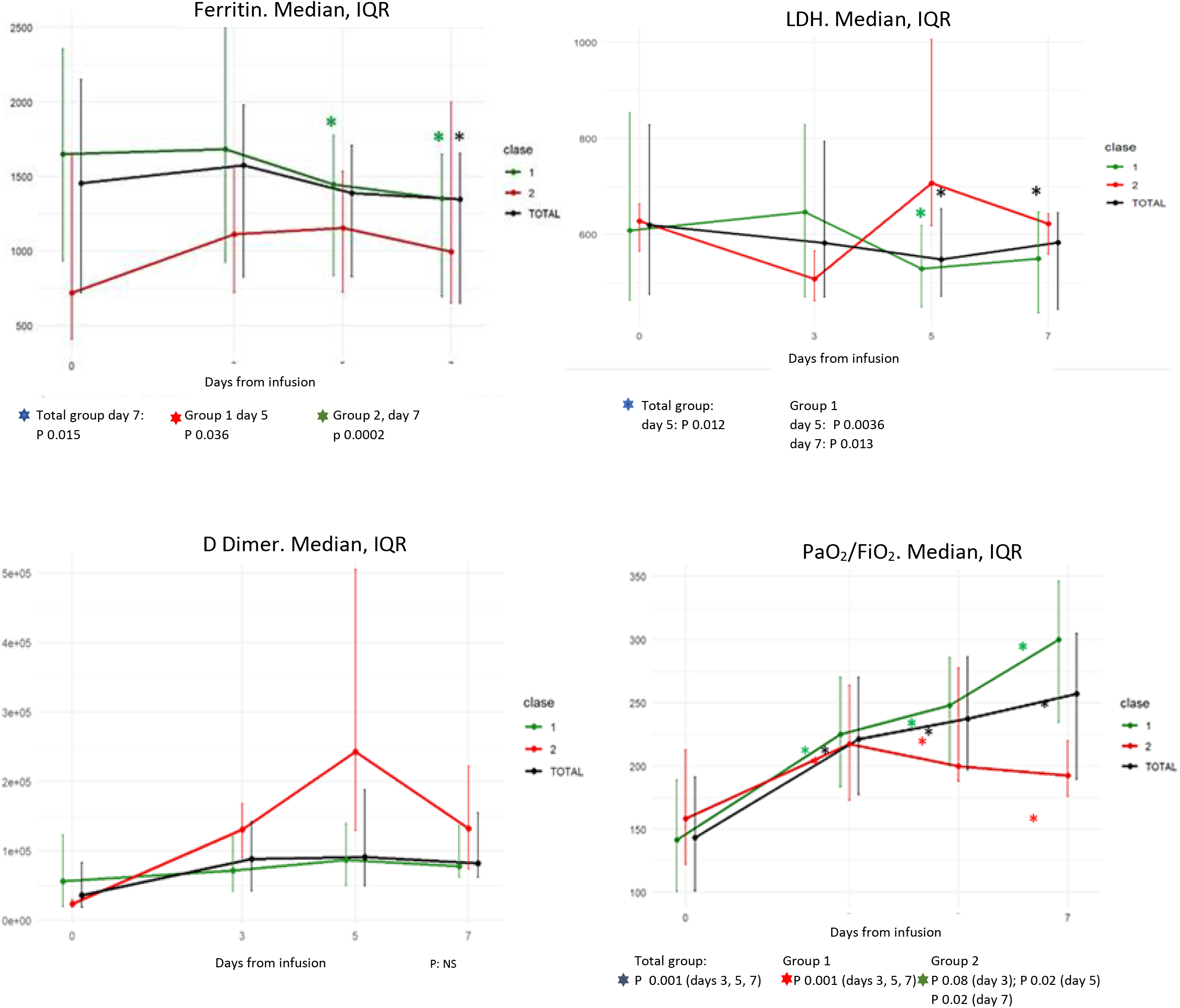
Evolution of inflammatory and respiratory laboratory parameters from infusion to days 3, 5 and 7

At the time of this report, from 87 patients, 22 died, 17 before 28 days post-infusion and 5 later on. From the 65 survivors, 49 had been discharged from the hospital, 16 remained hospitalized after day 28 (13 of them recovering in the general ward, 3 just weaned from mechanical ventilation) (fig. 4)

**Figure 4.**
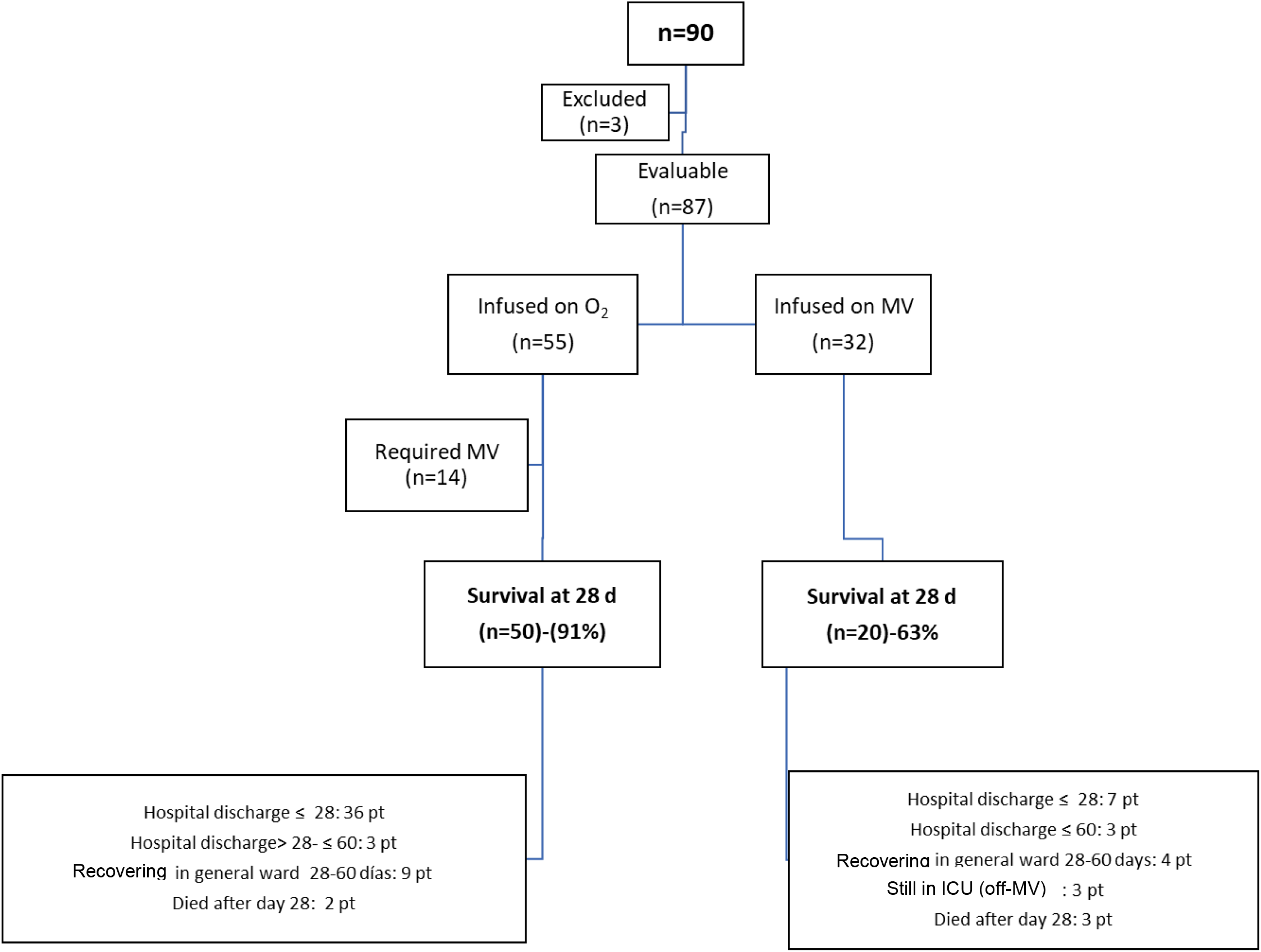
Flow chart of 90 severely ill COVID-19 patients and outcome after convalescent plasma infusion

## Discussion

This study represents to our knowledge the first report from South America on the feasibility and potential efficacy of convalescent plasma infusion therapy in SARS-CoV-2 infection. We report the observations of 87 evaluable patients with severe and / or critical pneumonia.

In this population, males and certain comorbidities, like diabetes, hypertension, and morbid obesity, were highly represented as reported in several series ^10^,^11^. Obesity rates registered in our population were comparable to that reported in patients hospitalized in intensive care units in the United States, which highlights this condition as a risk factor for developing severe forms of the disease and affecting people younger than initially reported ^12^. Twenty-three per cent of these seriously ill patients infused with plasma related to healthcare workers, highlighting their degree of exposure. The infection rate of health-care workers affected by the pandemic in Europe has been reported between 6 and 44%, depending on multiple factors such as the geographic zone and degree of exposure ^13^.

The safety of plasma infusion has recently been reported in 20,000 patients in whom foreseeable immediate adverse effects (circulatory overload, acute lung injury, and allergic reactions) were observed in less than 1% of cases ^6^. In the 112 infusions in our series, 1 episode of circulatory overload and a febrile reaction were reported, without the need to halt the infusion.

The overall mortality observed was 20% at 28 days, with a significantly lower rate in subjects who received the plasma infusion under spontaneous respiration with O_2_ supplementation compared to patients who received the infusion during mechanical ventilation (9% vs 37% respectively).

Patients infused with O_2_ supplements, despite high requirements provided with a NRO^2^-mask in most cases, progressed to intubation in 25%. In studies of patients with COVID-19 and respiratory failure with high O_2_ supplementation, the probability of requiring mechanical ventilation is high. In 2 Hospitals in New York, with 1150 hospitalized adults, 62% of the individuals with high O_2_ requirements (the majority with NRO^2^-mask) required intubation and mechanical ventilation ^10^. In the Recovery study, among patients included with O_2_ supplementation, the 28-day mortality was 26% in the control arm and 23% in the dexamethasone arm. Death rate and/or intubation was 32% and 28%, respectively, in that analysis period ^3^. The ICNARC report describes the evolution of 10,228 patients with COVID pneumonia. In 2591 patients with basic respiratory support, which includes an O_2_ mask with FiO_2_ equal to or greater than 50%, CPAP or non-invasive ventilation, mortality was 19.5% ^14^.

In our study, the mortality of patients on mechanical ventilation was 37%. Most of them were critically ill, met Berlin criteria of severe ARDS in 25%, with ventilation in the prone position in 67%, and with acute renal failure requiring hemodialysis in 10%. The death rate in patients on mechanical ventilation due to COVID19 pneumonia has been high in other series, reaching 88% in hospitals in a New York area ^15^. In our country there are still not enough data available, but in a preliminary report of an Argentine multicenter group, mortality in 47 mechanically ventilated patients was 62%. If only patients whose evolution was known were considered (since 8 remained alive, but on mechanical ventilation), the reported death rate reached 78% ^16^.

Convalescent plasma has been used as a treatment for numerous viral infections. The paradigmatic experience was the one that demonstrated its efficacy in AHF, produced by the Junín virus. In a randomized study against normal plasma, a significant reduction in lethality from 16.5% to 1.1% was observed ^4^. Regarding COVID-19 infection, after the first report of 5 patients infused in a hospital in China ^5^, reports from various countries were added. In a Seattle study of 20 infused patients versus 20 controls, the outcome was reported within 14 days. Only 6 of the patients were on mechanical ventilation at the time of infusion, 4 remained ventilated and 9 of each group were discharged within that period of analysis^17^. Another study of 115 plasma-infused cases compared to 74 controls reported a hospital discharge rate of 98% versus 78% respectively. Patients on mechanical ventilation and those with a high O_2_ requirement had been excluded ^18^. In a multicenter study from Wuhan, China, 103 patients were assigned to plasma versus control group, and no differences were observed in survival rates at 28 days, but clinical improvement was observed in subjects who received plasma (52% vs. 43%, P: ns). However, this difference was significant in patients with severe disease (hypoxemia without mechanical ventilation) since clinical improvement on the WHO scale was observed in 91% of treated patients vs. 68% of the control group (P = 0.03). A higher negative rate of viral RNA from respiratory secretions was also observed (87% vs. 37% at 72 h). This study was concluded before the planned end due to lack of recruitment and it is worth mentioning that the median to infusion from the onset of symptoms was 30 days ^19^. In another study of cases (n = 39) and controls (n = 156), an improvement in oxygen therapy requirements and a greater probability of survival was reported at the expense of the spontaneously breathing patients with O_2_^20^. A Dutch study with 86 patients compared plasma vs. controls. Anti-SARS-CoV-2 antibodies were detected before plasma infusion. Prognostic variables, hospital stay or clinical severity score at 15 days did not provide differences in mortality between arms ^21^. Finally, a study with 32 patients infused under spontaneous breathing, the intubation rate was 15.6% and the 30-day mortality was 22.5%, versus 34% observed from patient records from the same institution who did not receive plasma therapy. For the group of mechanically ventilated subjects, the mortality rate in those who received plasma was 46.7% and the comparative group from the same institution was 68.5% ^22^.

In our series, most of the infused patients (70%), improved at least 1 point of the score at 7 days and 62% at least 2 points at 14 days. At 7 days after plasma therapy, a significant improvement in PaO_2_ / FiO_2_ and ferritin level was observed.

Even steroids may improve survival in this setting (Recovery study ^3^), 49% of our patients did not receive them and had a good outcome in 68% of the cases.

There are several limitations of this study. The first one is that we do not have a control arm without convalescent plasma. When we designed it, there was no proven effective therapy for this disease. The mortality reported at the beginning of the pandemic in the most severe forms was high, and convalescent plasma was a possible strategy to apply in this situation (“*as much as possible, without stopping”* ^23^). The second limitation to consider is our unawareness of the neutralizing power of the infused plasma. We evaluated donor plasmas using IgG Ab against the nucleocapsid that did not establish neutralizing capacity, and in the first donors this evaluation was retrospective. When this test was available, the plasmas were selected by reading this method and most of the values were greater than 4 and never less than 3. According to some studies, the concordance between this serological response and viral neutralization suggests that a strong humoral response can be predictive of neutralizing activity, regardless of the selected target antigen ^8^. Furthermore, values greater than 4 had a correlation with a neutralizing titer of at least 1/320 ^24^.

Respiratory failure was the basic parameter to recruit patients in our study. In most cases it happened after several days of the onset of the initial symptoms. In a report of 4209 patients requiring admission to intensive care, the median time to onset of symptoms was 10 days ^25^. Although the most appropriate moment for infusion is still unknown and whether the main action of the plasma is viral neutralization, it can be assumed that the earlier, the better results could be obtained ^26^.

In conclusion, this report points out the feasibility and safety of convalescent plasma therapy, with an improvement in the clinical severity scale after the infusion. This beneficial effect is seen especially in the group of patients with severe COVID-19 pneumonia with O_2_ requirement. Understanding the limitation of a comparative analysis with the literature data, our observations allow us to speculate that the convalescent plasma could reduce the need to progress to intubation and thus have a positive impact on survival. Although we observed a lower survival rate in patients infused under mechanical ventilation, we cannot rule out even in these critical cases some positive action on survival. This assumption would require confirmation through randomized trials or inferred by careful studies with case-control analysis. Meanwhile, convalescent plasma administration is a valid and attractive option for the treatment of critically ill and seriously ill patients until other therapeutics can show to be effective and available.

## Data Availability

there are no supplementary materials

## Conflicts of Interest

The authors declare no conflict of interest related to the design of the study and its execution.

## Participant Institutions and members of the network

CEMIC, Sanatorio Güemes (Ignacio Romero), Hospital Fiorito (Viviana Falasco), Unión Obrera Metalúrgica (Fabián Romano), Sanatorio Itoiz (Mariana Chamadoira), Sanatorio Modelo de Caseros (Ana Cantillo), Sanatorio Franchín (Eleno Aquino), Sanatorio Monte Grande (Adrián Nuñez), Clínica Boedo (Orlando Campo), Sanatorio Bernal (Gonzalo Cortés), Hospital Abete de Malvinas Argentinas (Liliana Kumar), Clínica Modelo de Morón (Natalia Rondinelli), Clínica Calchaquí (Carlos Cremaschi), Hospital Iriarte de Quilmes (Gustavo Cañete), Hospital Argerich (Margarita Torres Boden), Clínica Bazterrica (Fernando Pálizas(^h^)), Cruz Blanca de Lanús (Marta Catalán), Sanatorio Adventista (Yael Pere) Hospital Eurnekián de Ezeiza (Luis Taco), Clínica Berazategui (Marilin Cavallín), Clínica Ranelagh (Nancy Soruco), Sanatorio Modelo de Quilmes (Omar Ada), Hospital HIGA Eva Perón de San Martín (María Carolina Salazar), Hospital Balestrini de la Matanza (Nydia Funes)

## Aknowledgments

Roberto Cacchione, MD, Guadalupe Carballa, MDl y Ac.Roberto Arana, MD (CEMIC)

*Servicio de Hemoterapia de CEMIC*, Technicians: Sergio Fridman, Analía Brest, Carla Farfán, Melody García Paredes, Natalia Goya, Lourdes Ilacqua, Carmen Kruppa, Roxana León Ruiz, Nidia López, Marcela Parrella, Claudia Quiroz, Marta Quiroz Fernández, Margarita Reta, Alejandra Sosa y Mariano Vázquez

*Instituto Universitario CEMIC*; Jimena Rey, MD; Mariano Wini, Tomás Rasines (students)

*Unidad de Investigación CEMIC:* Mónica Lombardo, MD; Victoria Marroquín, Stella Zarza

## Key Points

### Current Knowledge

High death rate has been reported for patients with severe COVID-19 pneumonia. Aside from positive effects of dexamethasone, especially for patients on mechanical ventilation, there are no other consolidate therapies. Earlier experiences with convalescent plasma infusion in viral diseases, including other coronavirus infections, lead to the testing of plasmatic therapy in COVID-19 disease.

Contribution of the paper to the current knowledge

This study of convalescent plasma infusion for patients with severe COViD-19 pneumonia, adds evidence concerning feasibility and safety of this therapy. It offers a potential efficacy, particularly in patients who receive the infusion before intubation and mechanical ventilation

## Notes

### Competing Interest Statement

The authors have declared no competing interest.

### Clinical Trial

NCT04535063

### Funding Statement

non funding

### Author Declarations

COMITE DE ETICA EN INVESTIGACION, CEMIC (CENTRO DE EDUCACION MEDICA E INVESTIGACIONES CLINICAS) Protocol Code: CEM-COV-19-01

## References

1. https://www.who.int/docs/default-source/coronaviruse/situation-reports/20200809-covid-19-sitrep-202.pdf?sfvrsn=2c7459f6_2

2. Berlin DA, Gulick RM, Martinez FJ. Severe Covid-19. N Engl J Med. 2020;10.1056/NEJMcp2009575.

3. RECOVERY Collaborative Group, Horby P, Lim WS, et al. Dexamethasone in Hospitalized Patients with Covid-19 – Preliminary Report [published online ahead of print, 2020 Jul 17]. N Engl J Med. 2020;10.1056/NEJMoa2021436.

4. Maiztegui JI, Fernandez NJ, de Damilano AJ. Efficacy of immune plasma in treatment of argentine haemorrhagic fever and association between treatment and a late neurological syndrome. Lancet. 1979;2(8154):1216-1217.

5. Shen C; Wang Z, Zhao F, et al. Treatment of 5 Critically Ill Patients With COVID-19 With Convalescent Plasma. JAMA. 2020; 323(16):1582-1589.

6. Joyner MJ, Bruno KA, Klassen SA, et al. Safety Update: COVID-19 Convalescent Plasma in 20,000 Hospitalized Patients. Mayo Clinic Proceedings. June 17,2020

7. https://www.uscovidplasma.org/

8. Jääskeläinen AJ, Kuivanen S, Kekäläinen E, et al. Performance of six SARS-CoV-2 immunoassays in comparison with microneutralization. J Clin Virol. 2020; 129:104512.

9. WHO Working Group on the Clinical Characterisation and Management of COVID-19 infection. A minimal common outcome measure set for COVID-19 clinical research. Lancet Infect Dis. 2020;20(8): e192-e197.

10. Cummings MJ, Baldwin MR, Abrams D, et al. Epidemiology, clinical course, and outcomes of critically ill adults with COVID-19 in New York City: a prospective cohort study. Lancet. 2020;395(10239):1763-1770.

11. Docherty AB, Harrison EM, Green CA, et al. Features of 20 133 UK patients in hospital with covid-19 using the ISARIC WHO Clinical Characterization Protocol: prospective observational cohort study BMJ 2020; 369:m1985

12. Kass DA, Duggal P, Cingolani O. Obesity could shift severe COVID-19 disease to younger ages. Lancet. 2020;395(10236):1544-1545.

13. Kursumovic E, Lennane S, Cook T M. Deaths in healthcare workers due to COVID-19: the need for robust data and analysis. Anaesthesia. 2020 Aug;75(8):989-992

14. https://www.icnarc.org/Our-Audit/Audits/Cmp/Reports

15. Richardson S, Hirsch JS, Narasimhan M, et al. Presenting Characteristics, Comorbidities, and Outcomes Among 5700 Patients Hospitalized With COVID-19 in the New York City Area [published online ahead of print, 2020 Apr 22] [published correction appears in doi: 10.1001/jama.2020.7681]. JAMA. 2020;323(20):2052-2059.

16. Plotnikov GA, Matesa A, Nadur JM et al. Rev Bras Ter Intensiva 2020; (Published ahead of print). http://rbti.org.br/imagebank/pdf/RBTI-0197-20-23.07.pdf.

17. Hegerova L, Gooley T, Sweerus KA, et al. Use of Convalescent Plasma in Hospitalized Patients with Covid-19 – Case Series [published online ahead of print, 2020 Jun 19]. Blood. 2020; blood.2020006964.

18. Abolghasemi H, Eshghi P, Cheraghali AM, et al. Clinical efficacy of convalescent plasma for treatment of COVID-19 infections: Results of a multicenter clinical study [published online ahead of print, 2020 Jul 15]. Transfus Apher Sci. 2020;102875.

19. Li L, Zhang W, Hu Y, et al. Effect of convalescent plasma therapy on time to clinical improvement in patients with Ssvere and life-threatening COVID-19: A randomized clinical trial JAMA. 2020;324(5):1-11.

20. Liu STH, Lin H-M, Baine I, et al. Convalescent plasma treatment of severe COVID-19: A matched 1 control study. MedRxiv 2020. 05. 20. 2020 102236; doi: https://doi.org/10.1101/2020.05.20.20102236

21. Gharbharan A, Jordans CE, Geurtsvankessel C, et al. Convalescent Plasma for COVID-19. A randomized clinical trial. medRxiv 2020.07.01.20139857; doi: https://doi.org/10.1101/2020.07.01.20139857

22. Donato Mi, Park S, Baker M, et al. Clinical and laboratory evaluation of patients with SARS-CoV-2 pneumonia treated with high-titer convalescent plasma: a prospective study. https://www.medrxiv.org/content/10.1101/2020.07.20.20156398v3. doi: https://doi.org/10.1101/2020.07.20.20156398.

23. Rubin R. Testing an Old Therapy Against a New Disease: Convalescent Plasma for COVID-19 [published online ahead of print, 2020 Apr 30]. JAMA. 2020;10.1001/jama.2020.7456. doi:10.1001/jama.2020.7456

24. Meschi S, Colavita F, Bordi L, et al. Performance evaluation of Abbott ARCHITECT SARS-CoV-2 IgG immunoassay in comparison with indirect immunofluorescence and virus microneutralization test [published online ahead of print, 2020 Jul 6]. J Clin Virol. 2020; 129:104539.

25. Grasselli G, Greco M, Zanella A, et al. Risk factors associated with mortality among patients with COVID-19 in intensive care Units in Lombardy, Italy [published online ahead of print, 2020 Jul 15]. JAMA Intern Med. 2020; e203539.

26. Joyner MJ, Senefeld JW, Klassen SA, et al. Effect of Convalescent Plasma on Mortality among Hospitalized Patients with COVID-19: Initial Three-Month Experience. MedRxiv 2020. 08.12. 20169359; doi: https://doi.org/10.1101/2020.08.12.20169359

